# Hidden burden of surgical cystic echinococcosis in Arequipa, Peru: An unrecognized population at risk

**DOI:** 10.1101/2025.08.01.25332595

**Authors:** Ricardo Castillo-Neyra, Wilfredo Gutierrez Zárate, María Paula Requena-Herrera, Lizzie Ortiz-Cam, Guillermo Porras, Jesus Marcelo Manturano Lopez, Jose Fernando Macedo Milla, Jorge L. Cañari-Casaño, Javier A. Bustos

## Abstract

Peru is endemic for *Echinococcus granulosus sensu lato (s.l.)*, a cestode that causes cystic echinococcosis (CE) in humans. Epidemiological and clinical research has focused extensively on the central and southern highlands, where the parasite is hyperendemic. However, the region of Arequipa, with active transmission of the parasite, has received much less attention. In 2011, a state clinical-surgical program for CE patients was launched in the city of Arequipa. Over less than two years, before program funds were reallocated, surgeons treated 177 patients. This volume underscores the urgent need to prioritize and include Arequipa in CE control programs in Peru. This study characterizes patients who underwent surgical treatment during the program’s duration and investigates evidence of active *E. granulosus s.l.* transmission in Arequipa. We extracted socio-demographic, epidemiological, and clinical data from 129 records of CE patients treated between December 2011 and November 2013. We compared frequencies across variables, including the patient’s age, sex, residence, education level, and occupation. We also mapped patient origins, with 40.3% being autochthonous to the Arequipa region. Patients’ median age was 26 (IQR: 16-45), 55% were men, and 69% were students, farmers, or housemakers. In contrast to other reports, where hepatic cases are predominant, 70% of the cases were pulmonary. The southern highlands of Peru, including Arequipa, suffer from endemic transmission of *E. granulosus s.l.* Limited access to healthcare hinders timely diagnosis and treatment, exacerbating the public health burden. While implementing or reinstating clinical-surgical units in Arequipa and other endemic areas would improve care and case monitoring, these measures alone are not sufficient. Effective control requires integrated One Health approaches and intersectoral collaboration across human, animal, and environmental health sectors.

**Author summary:** Cystic echinococcosis (CE) is a neglected parasitic disease caused by *Echinococcus granulosus sensu lato* (*s.l.*), which significantly impacts human and animal health in endemic regions, including southern Peru. This study analyzed medical records of patients diagnosed with CE at a major hospital in Arequipa, Peru, aiming to describe its epidemiology and clinical characteristics, and to highlight Arequipa as a historically overlooked region in CE research and control efforts. The findings reveal that CE disproportionately affects younger populations and rural communities, with men, students, farmers, and housemakers being at higher risk. Pulmonary involvement was notably more frequent than hepatic involvement, diverging from trends reported in other studies. The results underscore the importance of strengthening clinical-surgical units in the southern highlands and the necessity of including Arequipa in national control programs. Moreover, limited access to healthcare in rural and migrant populations underscores the importance of equitable resource allocation and sustained efforts to address health disparities. We emphasize the urgency of implementing integrated control strategies that combine human, animal, and environmental health approaches within a One Health framework. These findings provide valuable insights to inform policy and guide the development of cost-effective, interdisciplinary interventions focused on reducing the burden of CE in vulnerable populations.

## Introduction

Cystic echinococcosis (CE) is a zoonotic disease caused by the larval stage of *Echinococcus granulosus sensu lato (s.l.)*, a dog tapeworm [1]. Humans exposed to *E. granulosus s.l.*’ eggs may become infected, developing CE primarily in the liver and lungs [2,3], with prevalence levels as high as 9.3% in Peru [4–6], potentially the most affected country in the Americas [7,8]. In Peru, infection rates in sheep range from 38% to 87%, while in dogs they range from 32% to 88% [4–6,9,10]. These endemic and hyperendemic regions experience high demand for surgical interventions, which place a significant strain on the healthcare system [6]. CE disproportionately affects impoverished communities, requiring considerable resources for diagnosis and treatment [6,11–13]. Precise estimation of prevalence and assessments of disease burden are rare but highly needed [14]; unfortunately, many challenges, including limited resources, methodological issues, and lack of data, prevent them [6,14,15]. Disparities in infection distribution, limited access to imaging services, and the absence of robust disease reporting systems collectively contribute to a significant underestimation of the actual prevalence [16,17].

The annual economic burden of CE in Peru is approximately $6.3 million [6,18]. Of this, $2.4 million is attributed to human infections, with surgical treatment accounting for one-third of the cost and lost productivity comprising the remainder [6]. Livestock production losses add another $3.8 million [6,18]. The estimated burden of Disability-Adjusted Life Years (DALYs) linked to surgical cases in Peru is 1,139 annually, comparable to the combined total for Argentina, Brazil, Uruguay, and Chile [18]. This DALY count related to surgical cases in Peru is comparable to the impact of other significant infections such as malaria and leishmaniasis [19,20]. However, these calculations might not fully encompass the full impact of reduced productivity during pre- and post-surgical periods, as well as the effects of underdiagnosis which is significant for neglected tropical diseases.

Although the Peruvian regions in the central and southern highlands are well-studied and benefit from extensive epidemiological research and control programs, other areas with potential active transmission— such as the region of Arequipa—remain underexplored. In 2011, a state results-based budgeting program (PpR) strengthened the capabilities of the surgeons’ team at the Honorio Delgado Hospital in the city of Arequipa to treat individuals suspected of having CE [21]. PpR, a public management strategy linking resources to measurable outcomes [22], funded surgical teams, antiparasitic treatments, and computed tomography scans for confirming diagnosis and enhancing pre-surgical evaluations. Between December 2011 and November 2013, this program allowed treating surgically 177 patients, before funds were reallocated. During this period, surgeons reported improved surgical outcomes, including reduced complications, shorter hospital stays, higher rates of resolved cases, and fewer recurrences. These surgically treated cases require further investigation to identify patterns that could inform policies and guide the epidemiology of CE in the region.

Our study characterizes the epidemiological and clinical profile of patients who underwent surgical treatment for CE in the city of Arequipa during the PpR-supported program and identifies whether transmission of *E. granulosus s.l.* is active in Arequipa. We analyzed the geographical distribution of cases and compared demographic and clinical characteristics across different regions of origin. Our study contributes to the knowledge base necessary for devising targeted strategies to mitigate the impact of CE in the neglected southern highlands of Peru. We emphasize the high burden of CE in Arequipa, Peru, and the likelihood of numerous autochthonous cases in an area where CE has not been recognized officially as endemic, and where active transmission of *E. granulosus s.l.* is presumed to be non-existent.

## Methods

### Ethical considerations

Ethical approval was obtained from the International Review Boards from the University of Pennsylvania, Philadelphia, USA (approval number: 855517) and A.B. PRISMA, Lima, Peru (approval number: CE.0525.23).

### Study design

This retrospective study analyzed data from surgical CE patients in the largest hospital in the city of Arequipa between December 2011 and November 2013, during the duration of the PpR program.

### Study site and population

This study is situated in Arequipa, Peru’s second-largest city, which represents 4.9% of the national land area, located at an elevation of 2,328 meters above sea level [23]. As of 2013, the city had a population of 1.26 million [24]. Arequipa, located in southern Peru, encompasses both coastal and highland regions, with the Andes mountains running through it from north to south. Its weather is diverse, with temperatures ranging from 12°C to 29°C, as well as alternating rainy and dry seasons [25].

The study focused on individuals who underwent surgery for cystic echinococcosis between December 2011 and November 2013 in a regional hospital in Arequipa, Peru. Cases involving pulmonary, hepatic, and multiorgan cystic echinococcosis, where one of the affected organs was either the lung or liver, were included, while other locations were excluded for this analysis.

### Data collection

A search was conducted in the hospital’s Statistics Department database, applying a filter to select patients who had been hospitalized and received care by the surgery department. From this selection, we identified patients with a registered diagnosis of cystic echinococcosis using the relevant 10th edition International Classification of Diseases (ICD-10) codes (B67.0-B67.9). Then, we selected medical records that met the inclusion criteria and retrieved them from the hospital’s archive. Socio-demographic, epidemiological, and clinical information were collected using a RedCAP survey by two researchers and then cross-checked. Variables extracted included age, sex, educational level, occupation, geographic origin, cyst location, and recurrence associated with each case. In this study, we consider recurrence according to WHO guidelines, defined as the appearance of an active CE cyst (stages CE1–CE3) in the same location where a previously treated cyst was located, regardless of the type of treatment received [26].

### Data analysis

For statistical analysis and data visualizations, we used R software, version 4.4.2 [27] . Descriptive statistics, including median with interquartile range (IQR) and frequencies, were used to evaluate the distribution of socio-demographic and clinical variables. Additionally, we conducted bivariate analyses to compare regions of origin with other variables, using the Kruskal-Wallis test, the Chi-square test, and Fisher’s exact test. We created a map illustrating the origins of patients, providing a visual representation of the case’s spatial distribution throughout southern Peru.

## Results

From the 177 surgically treated CE cases reported in the system during the PpR program, we recollected the data from 166 medical records of patients who had an ICD-10 code for CE registered by the surgical department during their hospitalization. From this, we excluded twenty-three cases that received non-surgical treatment, nine suspected CE cases that were subsequently ruled out, three incomplete medical records, and two CE cases outside the liver or lung (vertebral and brain CE). Finally, 129 surgical CE cases were included in this study (Fig 1).

**Fig 1.**
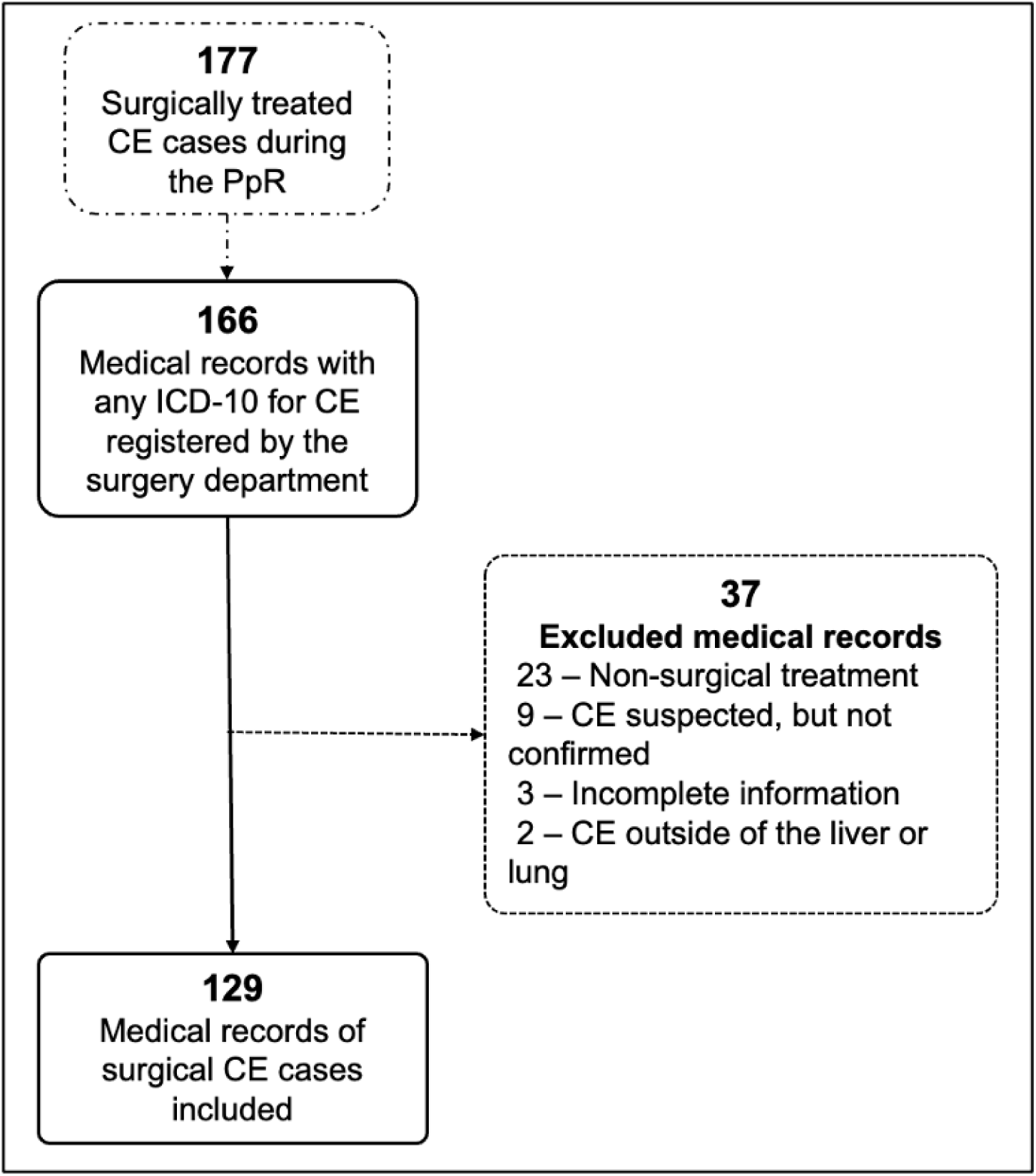
Flowchart of included/excluded medical records.

We stratified CE cases by geographical origin as Arequipa, Puno, and Others, the latter grouping those from Cusco, Moquegua, and Tacna (Table 1). The median age was 26 years (IQR: 16-45), and patients from Arequipa were older than those from Puno and Others (29, 23, and 22 years, respectively), but this difference was not statistically significant (p = 0.608). Patients from Arequipa and Others were predominantly males (58% and 62% respectively), while in Puno they represented only 49%, with no significant differences (p = 0.512). Secondary school was the most common educational attainment level (Arequipa: 44%, Puno: 51%, Others: 46%), followed by primary school (Arequipa: 31%, Puno: 22%, Others: 42%).

**Table 1.** Sociodemographic characteristics of CE surgical patients according to geographical origin (N=129).

| Characteristics | Overall,<br>N = 129 | Arequipa,<br>N = 52 | Puno,<br>N = 51 | Others,<br>N = 26 | p* |
| --- | --- | --- | --- | --- | --- |
| Age, median [IQR] | 26 [16, 45] | 29 [19, 45] | 23 [16, 47] | 22 [12, 42] | 0.608<sup>a</sup> |
| Male, n (%) | 71 (55%) | 30 (58%) | 25 (49%) | 16 (62%) | 0.512<sup>b</sup> |
| Education level, n (%) |  |  |  |  |  |
| Illiterate | 8 (6.2%) | 4 (7.7%) | 4 (7.8%) | 0 (0%) | 0.535<sup>c</sup> |
| Primary | 38 (29%) | 16 (31%) | 11 (22%) | 11 (42%) |  |
| Secondary | 61 (47%) | 23 (44%) | 26 (51%) | 12 (46%) |  |
| Higher | 15 (12%) | 6 (12%) | 7 (14%) | 2 (7.7%) |  |
| Unknown | 7 (5.4%) | 3 (5.8%) | 3 (5.9%) | 1 (3.8%) |  |
| Occupation, n (%) |  |  |  |  |  |
| Student | 45 (35%) | 13 (25%) | 19 (37%) | 13 (50%) | 0.080<sup>c</sup> |
| Farmer | 23 (18%) | 5 (9.6%) | 14 (27%) | 4 (15%) |  |
| Homemaker | 21 (16%) | 11 (21%) | 8 (16%) | 2 (7.7%) |  |
| Construction workers | 11 (8.5%) | 8 (15%) | 2 (3.9%) | 1 (3.8%) |  |
| Other | 11 (8.5%) | 8 (15%) | 2 (3.9%) | 1 (3.8%) |  |
| Merchant | 8 (6.2%) | 2 (3.8%) | 3 (5.9%) | 3 (12%) |  |
| Driver | 7 (5.4%) | 3 (5.8%) | 2 (3.9%) | 2 (7.7%) |  |
| Unknown | 3 (2.3%) | 2 (3.8%) | 1 (2.0%) | 0 (0%) |  |
| Cyst location, n (%) |  |  |  |  |  |
| Pulmonary | 90 (70%) | 32 (62%) | 40 (78%) | 18 (69%) | 0.437<sup>c</sup> |
| Hepatic | 26 (20%) | 13 (25%) | 7 (14%) | 6 (23%) |  |
| Multiorgan** | 13 (10%) | 7 (13%) | 4 (7.8%) | 2 (7.7%) |  |
| Complicated cyst, n (%) | 51 (40%) | 21 (40%) | 24 (47%) | 6 (23%) | 0.124<sup>b</sup> |
| Surgery year, n (%) |  |  |  |  |  |
| 2011 | 9 (7.0%) | 5 (9.6%) | 3 (5.9%) | 1 (3.8%) | 0.803<sup>c</sup> |
| 2012 | 79 (61%) | 33 (63%) | 31 (61%) | 15 (58%) |  |
| 2013 | 41 (32%) | 14 (27%) | 17 (33%) | 10 (38%) |  |
| Recurrence, n (%) | 4 (3.1%) | 1 (1.9%) | 2 (3.9%) | 1 (3.8%) | 0.840<sup>c</sup> |

<sup>a</sup> Kruskal-Wallis Test, <sup>b</sup> chi<sup>2</sup>, <sup>c</sup> Fisher's exact test
\* p values do not include unknown data and were calculated between Arequipa, Puno, and Others groups
\*\* Multiorgan: 11 are pulmonary and hepatic, 1 pulmonary and diaphragmatic, 1 pulmonary and pancreatic

Regarding occupation, most patients were students (Arequipa: 25%, Puno: 37%, Others: 50%), followed by homemakers in Arequipa (21%), farmers in Puno (27%), and Others (15%). Additional classified occupations included construction workers (8.5%), merchants (6.2%), and drivers (5.4%). The “Other” (8.5%) category primarily included a small number of individuals working independently, as well as cooks, fishermen, and shoemakers. There was no statistical difference in occupational distribution according to origin (p=0.080) (Table 1).

Most cases in this study were pulmonary cysts, followed by hepatic and multiorgan involvement, but there were no statistical differences across organs or by patient’s origin (p=0.437). Most cysts were non-complicated (Arequipa: 60%, Puno: 53%, Others: 77%), with no significant variation between groups (p=0.124) (Table 1).

Surgery was performed primarily in 2012 (62%), followed by 2013 (30%) and 2011 (7.8%). Since the program began in December 2011, this year has had the lowest number of surgeries. Recurrence was reported in only 3.1% of the cases, with no statistical difference between Arequipa, Puno, and other regions (p=0.840) (Table 1). Additionally, among 45 patients who reported traveling, 36 indicated visiting endemic areas. Out of 24 patients based in Arequipa who reported trips, 20 traveled to endemic areas.

The age distribution of CE cases varied significantly depending on the location of the cyst (Fig 2). Lung CE cases predominantly affected younger individuals, with a median of 22.5 years and a mean of 27.2 years, showing a right-skewed pattern. Hepatic CE cases, in contrast, were more frequent in middle-aged populations, with a median of 44.5 years and a mean of 44.1 years, reflecting a more symmetrical distribution. Multi-organ CE cases primarily occurred in slightly younger individuals, with a median of 20 years and a mean of 26.2 years, showing a right-skewed pattern.

**Fig 2.**
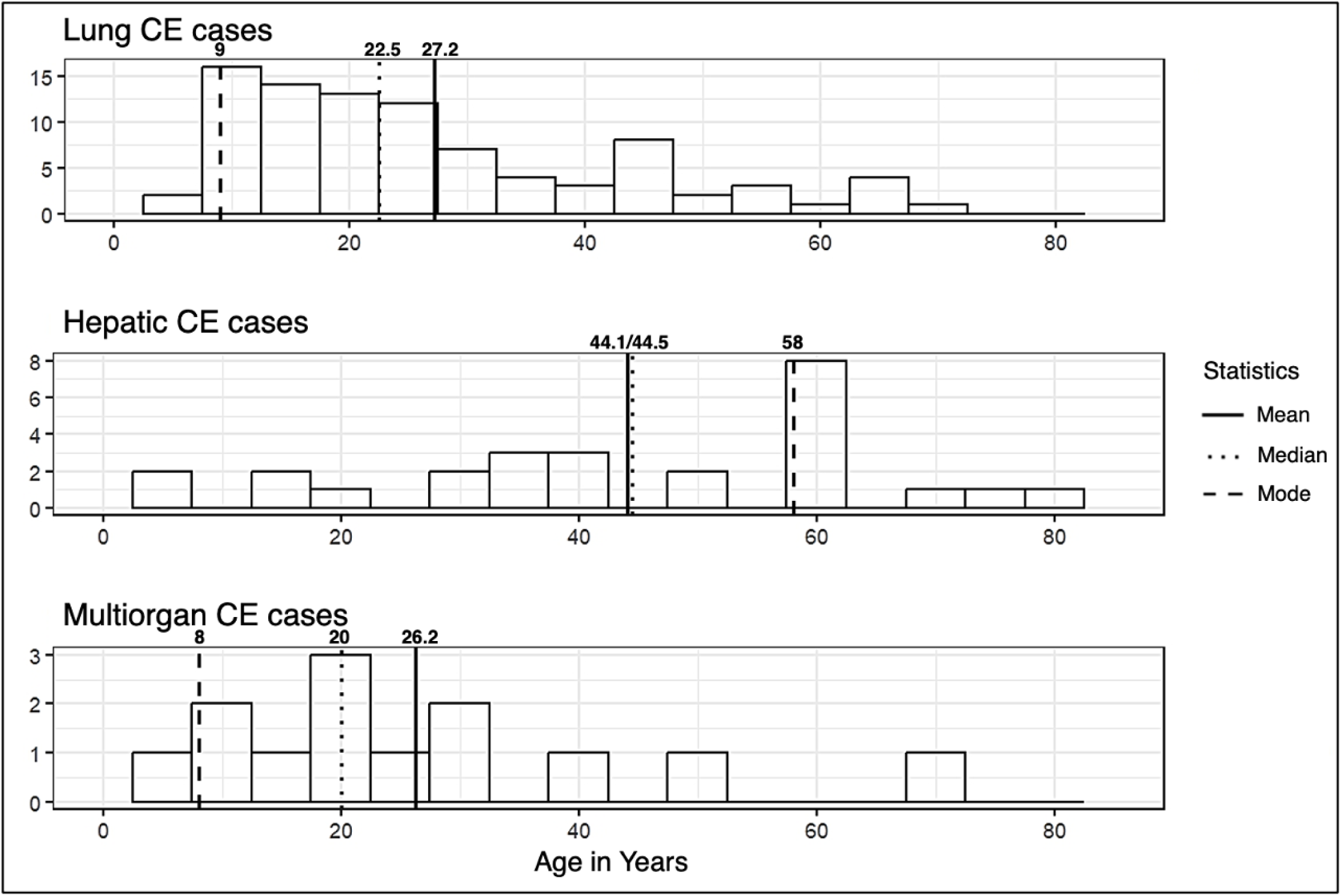
Age distribution of CE cases in Arequipa, Peru, from 2011 to 2013 according to cyst location. There was a higher concentration of cases in Arequipa, particularly in the Arequipa province, with the highest recorded cases (21–25) (Fig. 3). Puno and Cusco followed, with several provinces showing moderate case numbers (6–20) (Fig. 3).

**Fig 3.**
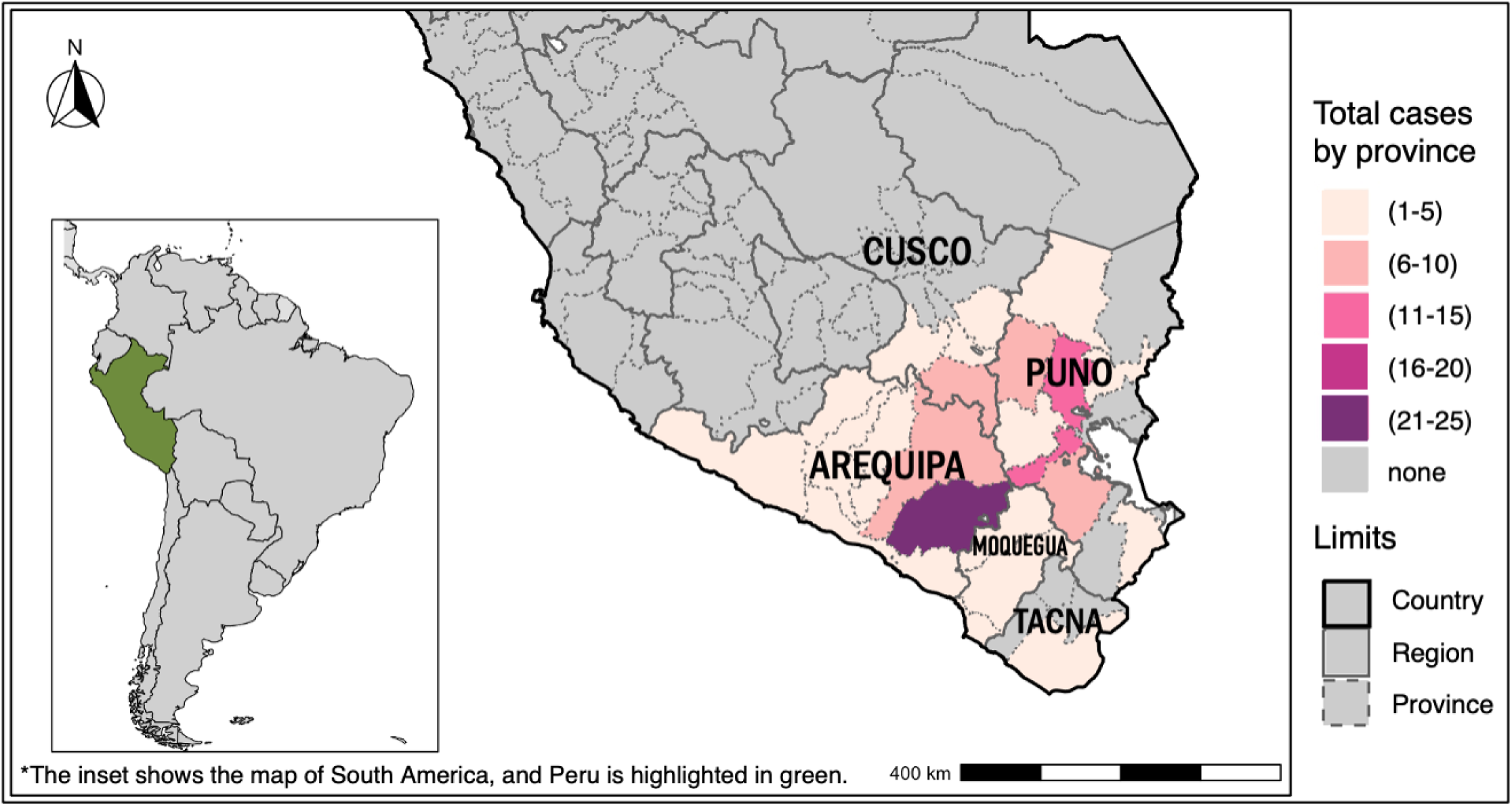
Geographical distribution of CE cases.

## Discussion

Arequipa is considered a non-endemic area for CE and has therefore received little attention from researchers and public health officials. However, in this study, we report a considerable number of CE cases, comparable to those in endemic regions of Peru, such as Puno [28–30], Cusco [31], Junín [32], and Huancayo [33,34]. The number of surgical cases observed in Arequipa was eight times higher than those reported in Huancayo, an endemic city, between 2000 and 2011(Santivanez, 2016). Notably, we identified 129 cases of surgically treated CE in an Arequipa hospital over a two-year period, with 40.3% of the cases being autochthonous. This number is relatively high compared to previously recognized endemic regions.

Very likely, the large number of cases treated in Arequipa is the result of the PpR program. Between 1970 and 1988, 95 pediatric patients, with an average age of 8.5 years, suffering from pulmonary and/or hepatic CE were examined at the Honorio Delgado Hospital [35]. Similarly, records from the Goyeneche Hospital pediatrics department, spanning from January 1993 to December 2002, revealed 16 pediatric cases, with a mean annual incidence rate of 2.65 cases. Most cases (75%) originated from urban areas in Arequipa, with the remainder coming from Puno [36]. In a separate study, 111 patients were reported positive to CE between 1996 and 2000, and 183 patients were treated at the Honorio Delgado Regional Hospital from 2008 to 2009 [37].

Arequipa has experienced significant growth of 0.32 million since the period covered by this study[38]. The city has consistently ranked among the top five most attractive destinations for internal migration, both in 2007 and 2017 [39]. The 2017 national census reflects this trend, reporting that 26.3% of Arequipa’s population were migrants—a 29.3% increase since 2007. Most migrants in Arequipa come from Puno (37.8%) and Cusco (28.2%) [40], both of which are regions endemic to *E. granulosus s.l.*.

Additionally, our study found that autochthonous Arequipa cases of CE reported travel histories to these areas. These migration patterns and travel histories likely influence the regional distribution of *E. granulosus s.l.* infections and place additional pressure on healthcare services in urban centers, such as Arequipa, where both local and imported cases converge.

Our geographic distribution analysis revealed a higher concentration of CE cases in the highland regions, particularly in known endemic areas such as Puno and Cusco, but also in Arequipa. In contrast, coastal regions such as Moquegua and Tacna, as well as rainforest zones within Puno and Cusco, reported fewer cases. Endemic regions such as Puno, Cusco, Junín, and Huancayo are located in the highlands at altitudes exceeding 3,000 meters above sea level, suggesting that CE cases are primarily concentrated in these elevated areas. Over 80% of the Peruvian highlands are used for pastoralism because of their historical, cultural, and geographical significance [41]. Ecological conditions favor livestock grazing, supporting animals such as sheep, goats, camelids, and cattle [42]. Shepherd dogs, commonly used to manage these herds, play a key role in maintaining the transmission cycle of *E. granulosus s.l.*. These environmental and agricultural practices contribute to the establishment of the parasite’s life cycle. These same favorable conditions are present in multiple provinces of the Arequipa region.

The distribution of surgical CE cases by sex in our study showed a slight predominance in males. This finding contrasts with other studies, where females tend to be predominant [43–45]. Regarding age distribution, cases were most common in younger individuals. These findings align with those of Pineda-Reyes et al. in a recent study, suggesting that active transmission occurs early in life, considering the years of the latent period before symptoms appear[46]. Additionally, cases were frequently observed among students (35%), farmers (18%), and homemakers (16%), highlighting the need for targeted control interventions within these populations.

Pulmonary cysts were observed three times more frequently than hepatic cysts, contrasting with findings from other studies, where hepatic cases typically predominate [47,48]. This higher prevalence may be due to the faster growth of cysts and more severe symptoms when the lungs are affected, prompting patients to seek medical assistance sooner [49]. Additionally, pulmonary cases were predominantly observed in younger patients, as greater tissue elasticity in children leads to faster cyst growth [50]. Moreover, most pulmonary cases were from Puno and other regions, where the affected patients tended to be younger than those from Arequipa.

Peru, as part of the Regional Program for the Elimination of Cystic Echinococcosis/Hydatidosis 2022– 2029 coordinated by the Pan American Health Organization, implements targeted control measures focused on regular praziquantel deworming of dogs every 90 days in endemic areas, pilot sheep vaccination with the EG95 vaccine, surveillance in dogs and humans, and specific education, mainly to livestock producers [51,52]. Despite existing integrated national guidelines, the human and animal health sectors often operate separately. In this study, we demonstrate the importance of clinical-surgical units in the southern highlands of Peru, although these are not sufficient on their own. When reviewing control programs worldwide, De la Cruz and collaborators[53] highlight that integrated efforts between human and animal health are more effective than those that focus on a single sector. Using a One Health approach would lead to more effective outcomes in controlling *E. granulosus s.l.* in Peru.

There are important limitations to consider in this study. Medical records were physical, which complicated data retrieval and review. As is common in many hospitals that rely on paper-based systems, the documentation was not homogeneous. Cases were identified through an electronic database of record numbers, and corresponding files were manually retrieved; however, not all could be found, introducing potential selection bias. These factors may affect the generalizability of our findings and emphasize the need for enhanced medical record-keeping systems, particularly in resource-constrained settings.

Our findings reveal a significant burden of CE in Arequipa, a region traditionally considered non-endemic. The demographic profile of CE cases in this study—mainly young males, students, farmers, and homemakers—further suggests specific at-risk populations that require prioritization for intervention. The high number of surgical cases (177 during the two-year PpR, compared to around 1–2 surgeries per month in recent years), the predominance of pulmonary involvement, and the observed link between migration and transmission patterns emphasize the urgent need for targeted public health action. Given the notable case numbers and Arequipa’s role as a major destination for internal migration, these results highlight the importance of incorporating Arequipa in national CE control programs alongside historically recognized endemic regions. Strengthening surveillance systems, improving access to early diagnosis in children and young population, and implementing cost-effective prevention strategies in Arequipa could be crucial in reducing CE transmission and mitigating its impact in southern Peru. These efforts must be framed within a One Health approach, recognizing the interconnected roles of human, animal, and environmental health.

## Data Availability

The study data are public and available in Github: https://github.com/mariarequena17/data_clincal_cases_ce.git

## Funding

RCN was supported by the National Institute of Allergy and Infectious Diseases (grant no. K01AI139284). RCN, JB, and LOC were supported by the Fogarty International Center (grant no. D43TW012741). JB and MPRH were supported by the Fogarty International Center (grant no. D43TW001140). The funders had no role in study design, data collection and analysis, decision to publish, or preparation of the manuscript.

## Notes

### Competing Interest Statement

The authors have declared no competing interest.

